# Long-Term Immunogenicity Study of an Aluminum Phosphate-Adjuvanted Inactivated Enterovirus A71 Vaccine in Children: An Extension to a Phase 2 Study

**DOI:** 10.1101/2023.12.21.23300173

**Authors:** Nan-Chang Chiu, Chien-Yu Lin, Charles Chen, Hao-Yuan Cheng, Erh-Fang Hsieh, Luke Tzu-Chi Liu, Cheng-Hsun Chiu, Li-Min Huang

## Abstract

**Background:** EV-A71 causes hand, foot, and mouth disease with potentially fatal complications such as encephalitis and acue flaccid myelitis in infants and children. This study examined the long-term immunity conferred by EV71vac, an inactivated EV-A71 vaccine based on the B4 subgenotype adjuvanted with aluminum phosphate, in children from the age of > 2 months to < 6 years for up to 5 years after the first immunization.

**Methods:** A total of 227 subjects from age of 2 months to 6 years who had previously received either EV71vac or placebo in the phase 2 clinical study were enrolled. Subjects were split into age groups: 2 years to < 6 years (Group 2b), 6 months to < 2 years (Group 2c), and 2 months to < 6 months (Group 2d). Serum samples were taken periodically for up to five years after the first dose for immunogenicity against EV-A71.

**Results:** At year 5, the neutralizing antibody titers against B4 subgenotype remained high at 621.38 to 978.20, 841.40 to 1159.93, and 477.71 to 745.07 for Groups 2b, 2c, and 2d, respectively. Cross-neutralizing titers at year 5 were 99.14 to 444.30 and 341.94 to 998.20 against B5 and C4a subgenotypes, respectively. Nearly all subjects remained seroprotected at year five (95.8 to 100%). No long-term safety issues were reported.

**Conclusion:** This study shows that using the current dosing regimen, EV71vac conferred persistent immunity against EV-A71 for at least five years after the first vaccination in children from the age of two months to six years.

**Summary:** EV-A71 neutralizing antibody persisted at high level throughout five years post vaccination in children of two months to six years old.

## Introduction

Enterovirus is a diverse group of single-stranded, positive-sense RNA viruses that include important human pathogens such as rhinovirus, poliovirus, coxsackievirus A, and enterovirus A71 [1]. Enterovirus A71 (EV-A71) is a highly contagious and significant member of the enterovirus family, causing periodic outbreaks of hand, foot, and mouth disease (HFMD) in infants and children, particularly in tropical regions. In severe cases, it can lead to serious or fatal complications such as myocarditis, acute flaccid myelitis, aseptic meningitis, and encephalitis [2]. The case fatality rate for patients diagnosed with EV-A71-associated HFMD in the Asia-Pacific region ranges from less than 0.5% to 19% [3].

The epidemiology of EV-A71 in the Asia-Pacific region follows a cyclic pattern, with major outbreaks occurring in cycles of 3 to 4 years, especially during the summer [3]. Currently, there are no US Food and Drug Administration (FDA) or European Medicines Agency (EMA)-approved vaccines available for EV-A71. However, there are three licensed inactivated whole virus vaccines based on the C4a subgenotype in China, developed by the Chinese Academy of Medical Sciences (CAMS), Sinovac Biotech, and Beijing Vigoo [4]. The World Health Organization (WHO) has subsequently developed recommendations to ensure the quality, safety, and efficacy of inactivated EV-A71 vaccines, based on the previously approved vaccines [5].

EV71vac is a formalin-inactivated, aluminum phosphate-adjuvanted EV-A71 vaccine based on the B4 subgenotype. It is the first EV-A71 vaccine for use in children as young as two months of age and has been shown to provide cross-reactivity against B5, C4a, and C5 subgenotypes [6]. In a large phase 3 clinical trial involving approximately 3,000 participants from multiple regions, the vaccine demonstrated 100% efficacy (96.8% in the Poisson regression model) in its target population [7]. The vaccine has been approved for use in children aged two months to six years in Taiwan and is currently under regulatory process in Vietnam [8, 9]. The current study is an extension study follow-up to our phase 2 clinical study to investigate the long-term immunity conferred by EV71vac in children aged from two months to less than six years. Participants from the phase 2 trial were monitored for neutralizing antibody titers against EV-A71 for up to five years after the first vaccination by the following age groups: 2 to < 6 years, 6 months to < 2 years, and 2 to < 6 months.

## Methods

### Study design and participants

This study was an extension study to the phase 2 main study of the EV71vac, which was a double-blind, randomized, placebo-controlled study to evaluate the safety and immunogenicity of children aged two months to 11 years [4]. In the previous study, the subjects were grouped by age into four groups, of which three of the groups were followed up during this extension study: Group 2b (2 to < 6 years), Group 2c (6 months to < 2 years), and Group 2d (2 to < 6 months). Subjects had previously received either EV71vac at 1.25 μg (LD), 2.5 μg (MD), 5μg (HD) or placebo (phosphate buffer saline with adjuvant aluminum phosphate at 150 μg/0.5 mL) and did not receive any further treatment in this extension study. Inclusion criteria are subjects who have completed participation in the original phase 2 study Groups 2b, 2c, and 2d, and have received protocol-specified doses of EV71vac or placebo (total of 2 doses in Group 2b, and 3 doses for Groups 2c and 2d), and the subjects’ guardians are able to understand and sign the informed consent form. The first study visit was 4 years ± 60 days after the first vaccination for Group 2b, and 3 years ± 60 days after the first dose for Groups 2c and 2d. Subjects of Group 2b remained in the study for approximately 12 months and had 2 clinic visits; subjects of Groups 2c and 2d remained in the study for approximately 24 months and had 3 clinic visits. Immunogenicity against EV-A71 virus antigen at each visit was assessed.

The trial protocol and informed consent form were approved by the Taiwan Food and Drug Administration and the ethics committee of each investigation sites: National Taiwan University Research Ethics Committee, Chang Gung Medical Foundation Institutional Review Board, and MacKay Memorial Hospital Instituional Review Board. The trial was conducted in accordance with the principles of the Declaration of Helsinki and Good Clinical Practice. An independent data and safety monitoring board (DSMB) was established to monitor safety data and the trial conduct. The clinical trial is registered at ClinicalTrials.gov with the identifier: NCT04072276

### Procedure and outcomes

The primary objective was to evaluate the long-term antibody titers of EV7vac at 4 years and 5 years after the first dose vaccination for subjects at the age of 2 years to < 6 years (Group 2b), and 3 years to 5 years after the first dose vaccination for subjects at the age of 2 months to < 2 years (Group 2c and 2d). The primary endpoints were to evaluate the immunogenicity and serum neutralizing antibody titers (B4 subgenotype) in terms of:

- Geometric mean titer (GMT) of EV-A71 neutralizing antibody titers at 4 years and 5 years after the first dose of vaccination for part 2b; and 3 years to 5 years after the first dose of vaccination for part 2c and 2d.
- Seroprotection rate (defined as Neutralizing Antibody titer ≥1: 32) at 4 years and 5 years after the first dose of vaccination for part 2b; and 3 years to 5 years after the first dose of vaccination for parts 2c and 2d.

The secondary objective was to evaluate the potential of cross-protection of the EV71 vaccine. Thus, the secondary endpoints were the neutralizing antibody titers against the non-vaccine subgenotypes (B5 and C4a) through GMT of neutralizing antibody titers against the above subgenotypes.

### Laboratory methods

Neutralizing antibody levels against EV-A71 subgenotypes B4, B5, and C4a were assayed using a cytopathogenic effect assay (CPE) as previously described [6]. Serum samples were inactivated at 56°C for 30 minutes, serially diluted from 1:8, and mixed with equal volumes of 100 TCID_50_ of virus for 2 hours. Virus-sera mixture and cells were incubated at 37±0.5°C with 5% CO2 for 5 to 7 days. Virus neutralization titer of serum was determined as the reciprocal of the highest dilution capable of inhibiting 50% of the CPE.

### Statistical analysis

The population sets used in the study for statistical analysis were the following:

Intent-to-follow (ITF) population:

– The ITF population included all enrolled subjects for whom data are available.

According-To-protocol (ATP) population:

– The ATP population for analysis of immunogenicity included all evaluable subjects (i.e., those meeting all eligibility criteria and complied with the procedures defined in the protocol during the study) for whom data concerning immunogenicity endpoint measures are available.

Safety evaluation was performed on the ITF population. Analysis of efficacy endpoints was performed on ATP. The main conclusion was made on the ATP population analysis results of the primary endpoints.

All results are presented using descriptive statistics. Neutralizing antibody titers below the cut-off value were given the value of half the cut-off for calculation purposes. All results were presented using descriptive statistics. Kruskal-Wallis test with corrected Dunn’s multiple comparisons test was used to compare the means between groups (2-tailed, alpha=0.05). Seroprotection rate (SPR) was defined as neutralizing antibody titer ≥1: 32. 95% confidence interval values were calculated with a two-sided unpaired t-test for GMT and binomial distribution estimation for SPR. The analysis was performed using Prism 6.01 (GraphPad).

## Results

In total, 227 subjects were screened in this study; no subject was excluded via screening, resulting in 227 enrolled subjects, including 91 subjects in Part 2b, 76 subjects in Part 2c and 60 subjects in Part 2d (Figure 1). All 227 eligible subjects belonged to ITF population and ATP population. All except 8 subjects (2 in Part 2b, 1 in Part 2c, and 5 in Part 2d) completed the study. The demographic characteristics of the study population is summarized in Table 1.

**Figure 1.**
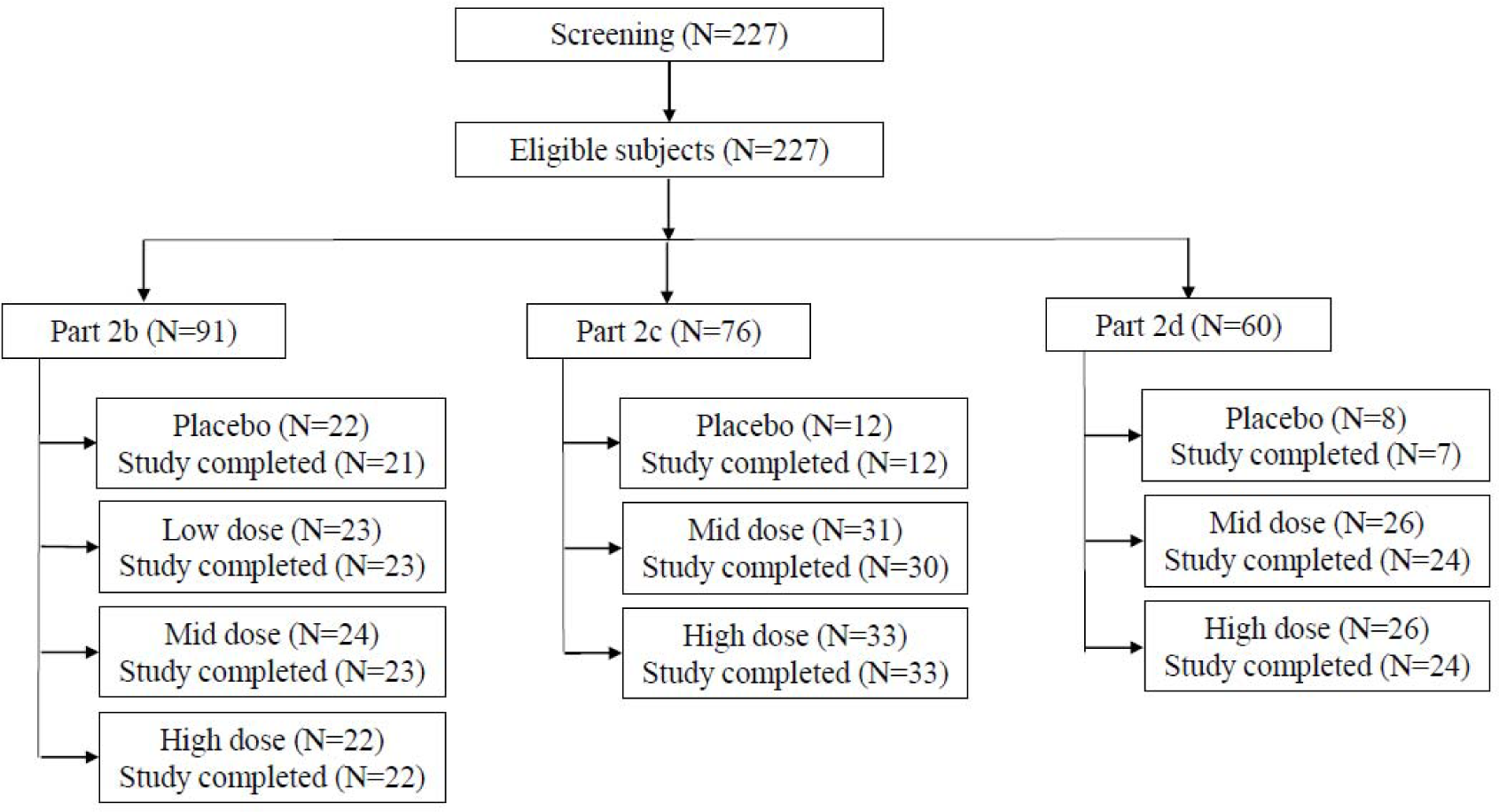
CONSORT flow diagram for the study.

**Table 1.** Summary of demographic characteristics of the participants at the time of screening* (ITF population)

|  | Group 2b<br>(2 – 6 years old at time of 1<sup>st</sup> dose) |  |  |  | Group 2c<br>(6 months – 2 years old at time of 1<sup>st</sup> dose) |  |  | Group 2d<br>(2 – 6 months old at time of 1<sup>st</sup> dose) |  |  |
| --- | --- | --- | --- | --- | --- | --- | --- | --- | --- | --- |
|  | Placebo | LD | MD | HD | Placebo | MD | HD | Placebo | MD | HD |
| N | 22 | 23 | 24 | 22 | 12 | 31 | 33 | 8 | 26 | 26 |
| Age (years)** | 8.5 (1.26) | 9.0 (1.14) | 8.2 (1.1) | 8.6 (1.1) | 5.5 (0.26) | 5.2 (0.36) | 5.2 (0.40) | 4.0 (0.16) | 4.1 (0.14) | 4.1 (0.14) |
| Male/Female (#) | 11/11 | 14/9 | 16/8 | 10/12 | 6/6 | 15/16 | 20/13 | 4/4 | 12/14 | 9/17 |
| Duration from first vaccination (years)** | 4.3 (0.07) | 4.4 (0.07) | 4.4 (0.07) | 4.2 (0.07) | 4.0 (0.09) | 3.9 (0.12) | 3.9 (0.12) | 3.7 (0.13) | 3.8 (0.14) | 3.8 (0.12) |

\* 4 years ± 60 days after the first vaccination for Group 2b, and 3 years ± 60 days after first dose for Groups 2c and 2d.
\*\*Data are expressed as mean (SD)

For Group 2b, at Year 4, the GMTs (95% CI) of EV71 B4 subgenotype-specific virus neutralizing antibody were 367.30 (95% CI 227.23 ∼ 593.70), 430.38 (258.53 ∼ 716.49), and 558.24 (359.16 ∼ 867.66) in subjects vaccinated with LD, MD, and HD, respectively (Figure 2 and Table 2). One year later at Year 5, the GMTs of the vaccinated remained high at 621.38 (398.89 ∼ 967.96), 978.20 (588.34 ∼ 1626.40), and 910.48 (549.55 ∼ 1508.47) for LD, MD, and HD groups, respectively. For Group 2c, at Year 3, the GMTs were 1206.99 (842.55 ∼ 1729.05) and 1576.81 (1171.27 ∼ 2122.77) for MD and HD, respectively, and further increased at Year 4 to 1653.18 (1178.10 ∼ 2319.83) and 2501.62 (1837.26 ∼ 3406.21) for MD and HD, respectively (Figure 2 and Table 2). However, at Year 5, the GMT values dropped to 841.40 (624.89 ∼ 1132.92) and 1159.93 (849.04 ∼ 1584.64) for MD and HD, respectively. A similar trend was observed in Group 2d, where the GMTs increased from 813.72 (538.80 ∼ 1228.92) to 1007.61 (609.16 ∼ 1666.71) for MD from Year 3 to Year 4, and from 956.17 (564.73 ∼ 1618.93) to 1699.14 (1141.13 ∼ 2530.02) for HD (Figure 2 and Table 2). At Year 5 the titers decreased to 477.71 (284.84 ∼ 801.17) and 745.07 (469.41 ∼ 1182.60) for MD and HD, respectively.

**Figure 2.**
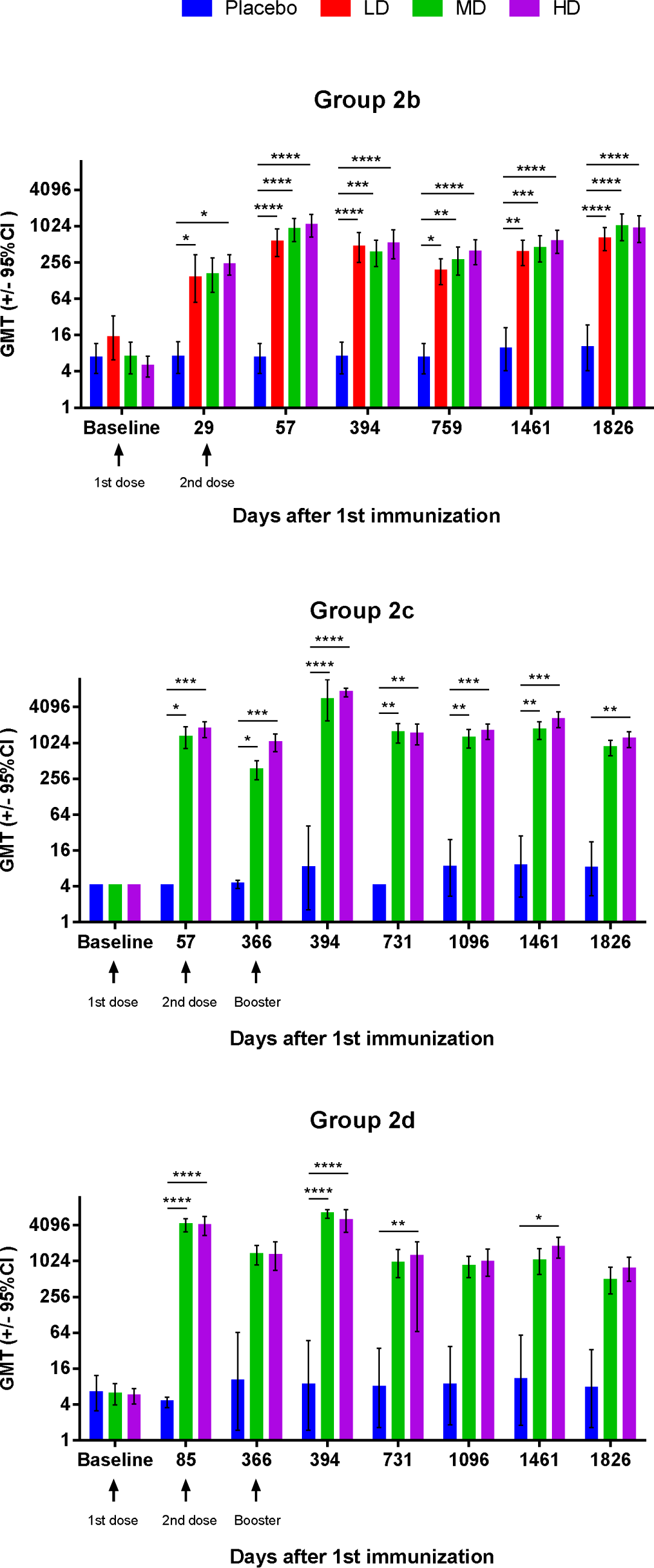
Immunogenicity of EV71vac during the extension study period. Neutralizing antibody titers for the B4 genotype were shown for Groups 2b, 2c, and 2d at various time points from the start of the main study (baseline) to five years after the first vaccination. The dose levels used were LD (1.25 μg), MD (2.5 μg), and HD (5 μg), and only Groups 2b received all three dose levels, whereas Groups 2c and 2d only received MD and HD. Arrows below the x-axis indicate the time of vaccination. Results are expressed as bars representing GMT and error bars representing a 95% confidence interval. Kruskal-Wallis test with corrected Dunn’s multiple comparisons test was used to compare the means between groups * = p < 0.05, ** = p < 0.01, *** = p < 0.001., **** = p < 0.0001.

**Table 2.** Neutralizing antibody titers against B4 subgenotype (ATP population)

|  | Group 2b |  |  |  | Group 2c |  |  | Group 2d |  |  |
| --- | --- | --- | --- | --- | --- | --- | --- | --- | --- | --- |
|  | Placebo | LD | MD | HD | Placebo | MD | HD | Placebo | MD | HD |
| Year 3 (Day 1096) |  |  |  |  |  |  |  |  |  |  |
| N | - | - | - | - | 12 | 31 | 33 | 8 | 26 | 26 |
| GMT (95% CI)* | - | - | - | - | 8.22 (2.75 – 24.56) | 1206.99 (842.55 – 1729.05) | 1576.81 (1171.27 – 2122.77) | 7.64 (1.65 – 35.28) | 813.72 (538.80 – 1228.92) | 956.17 (564.73 – 1618.93) |
| Year 4 (Day 1461) |  |  |  |  |  |  |  |  |  |  |
| N | 22 | 23 | 24 | 22 | 12 | 31 | 32 | 7 | 24 | 25 |
| GMT (95% CI)* | 9.25 (4.06 – 21.08) | 367.30 (227.23 – 593.70) | 430.38 (258.53 – 716.49) | 558.24 (359.16 – 867.66) | 8.69 (2.68 – 28.17) | 1653.18 (1178.10 – 2319.83) | 2501.62 (1837.26 – 3406.21) | 10.28 (1.80 – 58.92) | 1007.61 (609.16 – 1666.71) | 1699.14 (1141.13 – 2530.02) |
| Year 5 (Day 1826) |  |  |  |  |  |  |  |  |  |  |
| N | 21 | 23 | 23 | 22 | 12 | 30 | 33 | 7 | 24 | 24 |
| GMT (95% CI)* | 9.82 (4.09 – 23.57) | 621.38 (398.89 – 967.96) | 978.20 (588.34 – 1626.40) | 910.48 (549.55 – 1508.47) | 7.90 (2.78 – 22.43) | 841.40 (624.89 – 1132.92) | 1159.93 (849.04 – 1584.64) | 7.40 (1.64 – 33.23) | 477.71 (284.84 – 801.17) | 745.07 (469.41 – 1182.60) |

\*Geometric mean titer, 95% confidence interval calculated by two-sample t test

The dynamic of neutralizing antibody levels for all groups at each visit throughout the study is shown in Figure 2. Compared to the earlier stages of vaccination during the phase 2 main study, no significant increase or decrease was noted from 394 days after the first dose to the end of the study period (Figure 2 and Table S1). In all groups, neutralizing antibody titers peaked at four weeks after the final dose (2^nd^ dose for Group 2b, and booster dose for Groups 2c and 2d), and subsequently decreased but remain high until the fifth year (Figure 2).

In terms of seroprotection rate (SPR) against the B4 subgenotype, Groups 2b and 2c subjects remained completely seroprotected (100%) at Year 5 in all dose levels (Table 3). In Group 2d, one subject in the MD dose group had dropped enough to become not seroprotected, thus, resulting in an SPR of 95.8%, but the SPR of the HD group remained at 100% (Table 3). Interestingly, SPR in placebo groups increased during the course of the study to 14.3 – 19.0 % at Year 5 (Table 3).

**Table 3.**
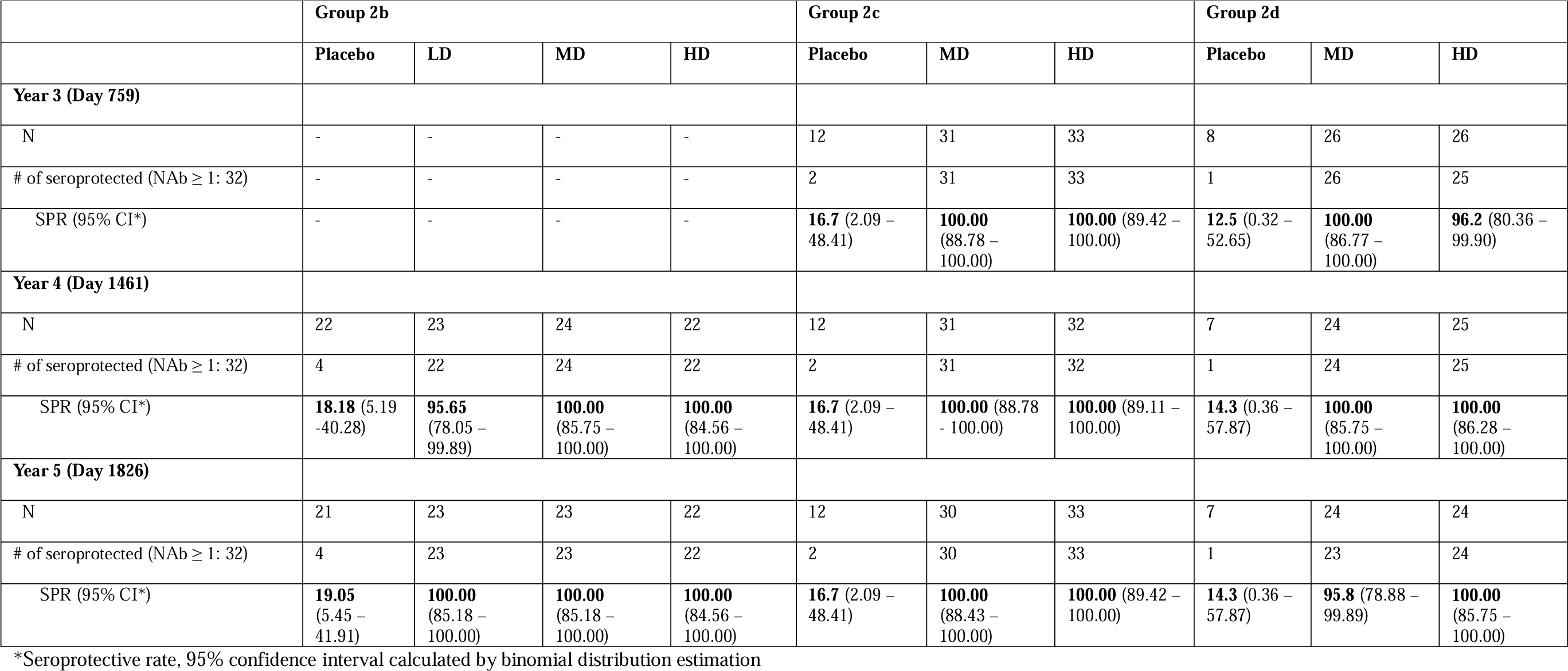
Seroprotection rate against B4 subgenotype (ATP population)

|  | Group 2b |  |  |  | Group 2c |  |  | Group 2d |  |  |
| --- | --- | --- | --- | --- | --- | --- | --- | --- | --- | --- |
|  | Placebo | LD | MD | HD | Placebo | MD | HD | Placebo | MD | HD |
| **Year 3 (Day 759)** |  |  |  |  |  |  |  |  |  |  |
| N | - | - | - | - | 12 | 31 | 33 | 8 | 26 | 26 |
| # of seroprotected (NAb ≥ 1: 32) | - | - | - | - | 2 | 31 | 33 | 1 | 26 | 25 |
| SPR (95% CI*) | - | - | - | - | **16.7** (2.09 – 48.41) | **100.00** (88.78 – 100.00) | **100.00** (89.42 – 100.00) | **12.5** (0.32 – 52.65) | **100.00** (86.77 – 100.00) | **96.2** (80.36 – 99.90) |
| **Year 4 (Day 1461)** |  |  |  |  |  |  |  |  |  |  |
| N | 22 | 23 | 24 | 22 | 12 | 31 | 32 | 7 | 24 | 25 |
| # of seroprotected (NAb ≥ 1: 32) | 4 | 22 | 24 | 22 | 2 | 31 | 32 | 1 | 24 | 25 |
| SPR (95% CI*) | **18.18** (5.19 – 40.28) | **95.65** (78.05 – 99.89) | **100.00** (85.75 – 100.00) | **100.00** (84.56 – 100.00) | **16.7** (2.09 – 48.41) | **100.00** (88.78 – 100.00) | **100.00** (89.11 – 100.00) | **14.3** (0.36 – 57.87) | **100.00** (85.75 – 100.00) | **100.00** (86.28 – 100.00) |
| **Year 5 (Day 1826)** |  |  |  |  |  |  |  |  |  |  |
| N | 21 | 23 | 23 | 22 | 12 | 30 | 33 | 7 | 24 | 24 |
| # of seroprotected (NAb ≥ 1: 32) | 4 | 23 | 23 | 22 | 2 | 30 | 33 | 1 | 23 | 24 |
| SPR (95% CI*) | **19.05** (5.45 – 41.91) | **100.00** (85.18 – 100.00) | **100.00** (85.18 – 100.00) | **100.00** (84.56 – 100.00) | **16.7** (2.09 – 48.41) | **100.00** (88.43 – 100.00) | **100.00** (89.42 – 100.00) | **14.3** (0.36 – 57.87) | **95.8** (78.88 – 99.89) | **100.00** (85.75 – 100.00) |

\*Seroprotective rate, 95% confidence interval calculated by binomial distribution estimation

Neutralizing antibody titers against B5 and C4a subgenotypes also remained high throughout the study period. At Year 5, the GMTs against B5 ranged from 156.77 to 242.78, 372.59 to 444.30, and 99.14 to 174.63 for Groups 2b, 2c, and 2d, respectively (Figure 3 and Table 4). At Year 5, the GMTs against C4a ranged from 341.94 to 436.72, 641.68 to 998.20, and 350.60 to 717.39 for Groups 2b, 2c, and 2d, respectively (Figure 3 and Table 5).

**Figure 3.**
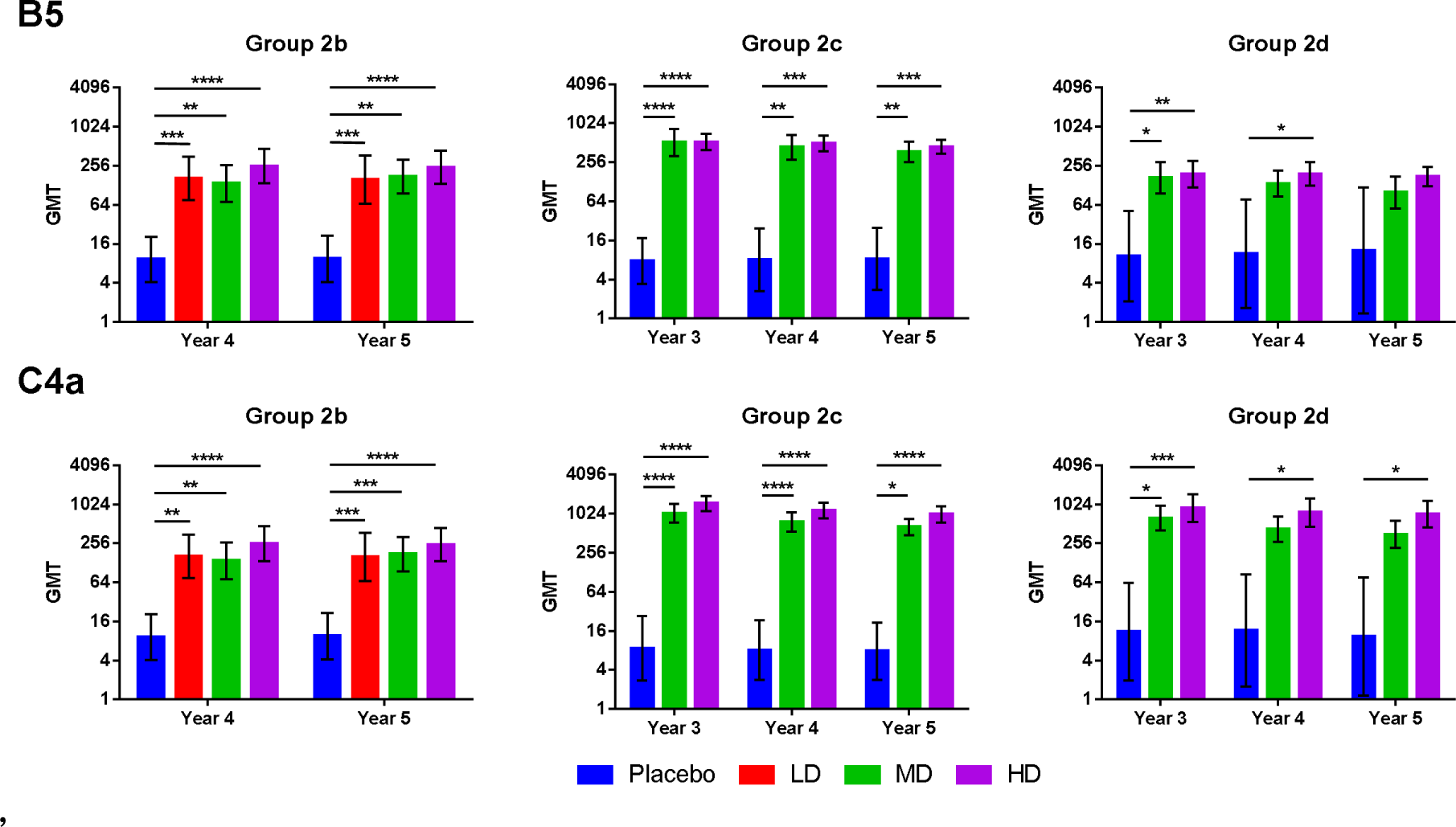
Immunogenicity of EV71vac during the extension study period. Neutralizing antibody titers for B5 and C4a genotypes were shown for Groups 2b, 2c, and 2d at three to five years after the first vaccination. The dose levels used were LD (1.25 μg), MD (2.5 μg), and HD (5 μg), and only Groups 2b received all three dose levels, whereas Groups 2c and 2d only received MD and HD. Results are expressed as bars representing GMT and error bars representing a 95% confidence interval. Kruskal-Wallis test with corrected Dunn’s multiple comparisons test was used to compare the means between groups * = p < 0.05, ** = p < 0.01, *** = p < 0.001., **** = p < 0.0001.

**Table 4.** Neutralizing antibody titers against B5 subgenotype (ATP population)

|  | Group 2b |  |  |  | Group 2c |  |  | Group 2d |  |  |
| --- | --- | --- | --- | --- | --- | --- | --- | --- | --- | --- |
|  | Placebo | LD | MD | HD | Placebo | MD | HD | Placebo | MD | HD |
| **Year 3 (Day 759)** |  |  |  |  |  |  |  |  |  |  |
| N | - | - | - | - | 12 | 31 | 33 | 8 | 26 | 26 |
| GMT (95% CI)* | - | - | - | - | **7.72** (3.40 – 17.51) | **520.84** (323.70 – 838.04) | **528.70** (395.11 – 707.44) | **10.33** (2.06 – 51.86) | **168.44** (96.61 – 293.69) | **190.33** (118.71 – 305.15) |
| **Year 4 (Day 1461)** |  |  |  |  |  |  |  |  |  |  |
| N | 22 | 23 | 24 | 22 | 12 | 31 | 32 | 7 | 24 | 25 |
| GMT (95% CI)* | **9.23** (4.11 – 20.76) | **162.61** (75.57 – 349.91) | **137.48** (71.40 – 264.70) | **253.57** (136.64 – 470.56) | **8.05** (2.66 – 24.41) | **438.25** (284.00 – 676.28) | **500.45** (381.76 – 656.05) | **11.26** (1.64 – 77.07) | **135.77** (86.14 – 213.98) | **192.58** (127.73 – 290.35) |
| **Year 5 (Day 1826)** |  |  |  |  |  |  |  |  |  |  |
| N | 21 | 23 | 23 | 22 | 12 | 30 | 33 | 7 | 24 | 24 |
| GMT (95% CI)* | **9.46** (4.12 – 21.70) | **156.77** (66.53 – 369.40) | **174.44** (95.38 – 319.05) | **242.78** (134.57 – 437.98) | **8.29** (2.75 – 24.96) | **372.59** (257.36 – 539.42) | **444.30** (345.73 – 570.96) | **12.68** (1.35 – 118.92) | **99.14** (56.14 – 175.05) | **174.63** (123.16 – 247.62) |

\*Geometric mean titer, 95% confidence interval calculated by two-sample t test

**Table 5.** Neutralizing antibody titers against C4a subgenotype (ATP population)

|  | Group 2b |  |  |  | Group 2c |  |  | Group 2d |  |  |
| --- | --- | --- | --- | --- | --- | --- | --- | --- | --- | --- |
|  | Placebo | LD | MD | HD | Placebo | MD | HD | Placebo | MD | HD |
| **Year 3 (Day 759)** | - |  |  |  |  |  |  |  |  |  |
| N | - | - | - | - | 12 | 31 | 33 | 8 | 26 | 26 |
| GMT (95% CI)* | - | - | - | - | **8.58** (2.73 – 26.93) | **1042.52** (744.02 – 1460.78) | **1470.06** (1117.95 – 1933.08) | **11.02** (1.95 – 62.26) | **629.22** (406.03 – 975.09) | **897.20** (545.70 – 1475.12) |
| **Year 4 (Day 1461)** |  |  |  |  |  |  |  |  |  |  |
| N | 22 | 23 | 24 | 22 | 12 | 31 | 32 | 7 | 24 | 25 |
| GMT (95% CI)* | **9.66** (4.12 – 22.64) | **317.04** (190.82 – 526.75) | **313.83** (205.42 – 479.47) | **575.40** (386.64 – 856.30) | **8.05** (2.80 – 23.19) | **769.43** (544.02 – 1088.23) | **1147.36** (871.77 – 1510.06) | **11.64** (1.57 – 86.00) | **426.00** (271.14 – 669.32) | **760.19** (461.95 – 1250.97) |
| **Year 5 (Day 1826)** |  |  |  |  |  |  |  |  |  |  |
| N | 21 | 23 | 23 | 22 | 12 | 30 | 33 | 7 | 24 | 24 |
| GMT (95% CI)* | **9.56** (4.12 – 22.16) | **341.94** (208.89 – 559.74) | **353.69** (222.40 – 562.47) | **436.72** (277.37 – 687.61) | **7.79** (2.84 – 21.37) | **641.68** (480.27 – 857.32) | **998.20** (742.61 – 1341.75) | **9.40** (1.16 – 76.09) | **350.60** (216.88 – 566.76) | **717.39** (447.75 – 1149.41) |

\*Geometric mean titer, 95% confidence interval calculated by two-sample t test

For long-term safety evaluation, physical examinations were performed at all visits. Overall, relative few subjects presented medical conditions s in both placebo and vaccine groups. As these medical conditions occurred years after vaccination, these were deemed less likelyto be related to the vaccine. No particular trends were observed between Placebo and each EV71 vaccine dose group (Table S2).

## Discussion

In this study, we investigated the long-term immunity for up to five years after the first dose of EV71vac in children aged from two months to six years. To our knowledge, this is the first study of long-term immunity imparted by the EV-A71 vaccine involving children aged from two to six months, which makes the findings significant in filling this knowledge gap.

Our data have shown that the antibody immunity elicited by EV71vac persisted throughout five years of study period against not only the B4 vaccine subgenotype, but also other subgenotypes. Clinical evidence from our phase 3 efficacy trial also suggested that EV71vac cross-protected against non-B4 subgenotypes [7]. The vaccine can therefore provide a strong foundation for protection against the EV-A71 through infancy and childhood. The finding is crucial, considering that the age group of two to six months is particularly vulnerable to severe complications of EV-A71 infection. In the previous phase III study, the number of subjects who reported adverse events was similar in EV71vac and placebo group during the 14-month observation period, and almost all reported solicited adverse events were mild and self-limited [7]. The results of this extension study further suggest that this EV71 vaccine is safe up to 5 years after the first dose as investigated by clinical visits. .

Our results are in line with the current knowledge regarding the durability of post-vaccination antibody levels, which show that antibody titers peaked at one month after the last vaccination, and remained stable at much higher levels than pre-vaccination [10]. It has been found that long-term antibody maintenance after exposure or vaccination is mainly attributed to: 1) Short-lived plasma cells are triggered by re-exposure to antigen which causes memory B cells to divide and differentiate; and 2) Long-lived plasma cells, which are derived from germinal center B cells and reside in bone marrows and secret high-affinity antibodies without re-exposure to antigens (LLPCs) [11, 12]. Although the increase in GMT and SPR could be attributed to the natural infection of EV-A71 during the course of the study, the SPR in our placebo groups suggested that it plays a minimal role if any. The increase of SPR in our placebo groups was 14.3% to 19.0% in the fifth year, noticeably lower than that observed in a five-year study in China, where SPR of the placebo group increased from 4.76% to 71.43% within 5 years [13]. For similar reason, the maintenance of immune persistence after vaccination is less likely due to exposure to EV-A71 virus in environment, which could strengthen the neutralizing antibody levels against EV-A71, as previously observed in studies in China [13–15]. One Chinese study has observed dramatic waning antibody titers just 6 months after the primary series of 2 vaccinations, from 509.0–1383.2 on day 56 to 58.8–177.4 at 8 months post-vaccination in all age and treatment groups [16]. In a subsequent study for the same vaccine as the booster, the same subjects showed that the neutralizing antibody titers became stabilized and slightly increased to 138.2-264.3 at the time of start of booster study (one year after the first dose) [17]. The authors also attributed to the slight increase of antibody titers post-vaccination to natural booster due to exposure to EV-A71 [17]. In contrast, our study did not observe any significant decrease in antibody titers up to five years after the first vaccination, implying that the antigen and adjuvant formulation/dosage is superior in maintaining the level of antibody titer through the study period (Figure 2).

The observed long-term immunity has important implications for public health and disease prevention. Children within the two to six months age range are at a critical stage of development, and providing them with adequate protection against infectious diseases is paramount. By demonstrating the vaccine’s ability to induce long-lasting immunity in this age group, we contribute valuable information to guide immunization strategies and policies for infants and young children. Large-scale EV-A71 outbreaks which occur in waves are attributed to a younger naïve population and thus, vaccination of this younger population is key to preventing and breaking the cycle of EV-A71 outbreaks [18]

We acknowledge the limitations of this study. The sample size was relatively small due to the selection of participants from the phase 2 trial. Long-term follow-up studies of efficacy in vaccinated individuals with larger cohorts could be conducted to validate and further investigate the real-world effectiveness of EV71vac against infection. The lack of immunogenicity data in the literature for the two to six months age group also limited our ability to compare our findings in this age group directly with previous studies. Further analysis is required to study the modeling of immune persistence such as half-life and decay of neutralization antibody response.

In conclusion, our study provides novel insights into the long-term immunity conferred by EV71vac in children aged from two months to six years, particularly in the vulnerable age group of two to six months which is not covered by the previously approved EV-A71 vaccines.

## Acknowledgements

We would like to thank all of the participants and study team members in the participating hospitals: National Taiwan University Hospital, Linkou Chang Gung Memorial Hospital, MacKay Memorial Hospital, and MacKay Memorial Hospital Hsinchu Branch. We would also like to thank fellow colleagues of Medigen Vaccine Biologics Corporation for reviewing and providing comments and input to the manuscript.

## Author contributions

H.-Y. C., E.-F. H., and L.-M. H. designed the study and experiments. N.-C. C., C.-Y. ., C.-H. C., and L.-M. H. were principal investigators and were responsible for acquisition and interpretation of data. L. T.-C. L. analyzed and visualized data, and drafted the manuscript. C. C. coordinated the project and provided administrative support. All authors have reviewed and approved of the final version of the manuscript.

## Data Availability

Data produced in the study are available upon reasonable request to the authors

