## Supplementary material for "Long-Term Immunogenicity Study of an Aluminum Phosphate-Adjuvanted Inactivated Enterovirus A71 Vaccine in Children: An Extension to a Phase 2 Study": Tables S1 and S2

Table S1. Neutralizing antibody titers and seroprotection rate against B4 subgenotype from early stages of vaccination (ATP population)

|  | **Group 2b** | | | | **Group 2c** | | | **Group 2d** | | |
| --- | --- | --- | --- | --- | --- | --- | --- | --- | --- | --- |
|  | **Placebo** | **LD** | **MD** | **HD** | **Placebo** | **MD** | **HD** | **Placebo** | **MD** | **HD** |
| **Baseline** | - | | | |  | | |  | | |
| N | 23 | 24 | 21 | 23 | 10 | 20 | 23 | 8 | 22 | 24 |
| GMT (95% CI*) | **6.56** (3.69 – 11.65) | **14.41** (6.19 – 33.55) | **6.71** (3.66 – 12.33) | **4.84** (3.26 -7.17) | **4.00** (4.00 – 4.00) | **4.00** (4.00 – 4.00) | **4.00** (4.00 – 4.00) | **6.22** (3.14 – 12.35) | **5.94** (3.96 – 8.91) | **5.49** (4.09 – 7.37) |
| # of SPR (NAb ≥ 1: 32) | 3 | 8 | 3 | 1 | 0 | 0 | 0 | 2 | 2 | 2 |
| Seroprotection rate (95% CI**) | **13.04** (2.78 – 33.59) | **33.33** (15.63 – 55.32) | **14.29** (3.05 – 36.34) | **4.35** (0.11 – 2.19) | **0.00** (0.00 – 30.85) | **0.00** (0.00 – 16.84) | **0.00** (0.00 – 14.82) | **25.00** (3.19 – 65.09) | **9.09** (1.12 – 29.16) | **8.33** (1.03 – 27.00) |
|  | **Prior to 2^nd^ dose (Day 29)** | | | | **28 days after 2^nd^ dose (Day 57)** | | | **28 days after 2^nd^ dose (Day 85)** | | |
| N | 23 | 24 | 21 | 23 | 10 | 20 | 23 | 8 | 22 | 24 |
| GMT (95% CI*) | **6.8** (3.7 – 12.5) | **138.59** (55.55 – 345.76) | **157.45** (82.22 – 301.51) | **231.54** (156.75 – 342.02) | **4.00** (4.00 – 4.00) | **1256.08** (830.57 – 1899.61) | **1713.27** (1267.82 – 2315.24) | **4.36** (3.55 – 5.35) | **4075.30** (3177.56 – 5226.66) | **3957.93** (2728.31 – 5741.72) |
| # of SPR (NAb ≥ 1: 32) | 3 | 18 | 20 | 23 | 0 | 20 | 23 | 0 | 22 | 24 |
| Seroprotection rate (95% CI**) | **13.04** (2.78 – 33.59) | **72.00** (50.61 – 87.93) | **95.24** (76.18 – 99.88) | **100.00** (85.18 – 100.00) | **0.00** (0.00 – 30.85) | **100.00** (83.16 – 100.00) | **100.00** (85.18 – 100.00) | **0.00** (0.00 – 36.94) | **100.00** (84.56 – 100.00) | **100.00** (85.75 – 100.00) |
|  | **28 days after 2^nd^ dose (Day 57)** | | | | **Prior to booster dose (Day 366)** | | | **Prior to booster dose (Day 366)** | | |
| N | 23 | 24 | 21 | 23 | 10 | 20 | 23 | 8 | 22 | 24 |
| GMT (95% CI*) | **6.56** (3.71 – 11.60) | **544.43** (320.86 – 923.78) | **885.16** (568.02 – 1379.38) | **1033.99** (668.37 – 1599.63) | **4.29** (3.67 – 5.02) | **358.57** (247.82 – 518.82) | **1026.85** (727.45 – 1449.48) | **9.76** (1.48 – 64.36) | **1281.18** (875.59 – 1874.66) | **1239.88** (713.02 – 2156.05) |
| # of SPR (NAb ≥ 1: 32) | 3 | 24 | 21 | 23 | 0 | 20 | 23 | 1 | 22 | 24 |
| Seroprotection rate (95% CI**) | **13.04** (2.78 – 33.59) | **100.00** (85.76 – 100.00) | **100.00** (83.89 – 100.00) | **100.00** (85.18 – 100.00) | **0.00** (0.00 – 30.85) | **100.00** (83.16 – 100.00) | **100.00** (85.18 – 100.00) | **12.50** (0.32 – 52.65) | **100.00** (84.56 – 100.00) | **100.00** (85.75 – 100.00) |
|  | **1 year after 2^nd^ dose (Day 394)** | | | | **28 days after booster dose (Day 394)** | | | **28 days after booster dose (Day 394)** | | |
| N | 23 | 24 | 21 | 23 | 10 | 20 | 23 | 8 | 22 | 24 |
| GMT (95% CI*) | **6.71** (3.66 – 12.29) | **449.44** (254.33 – 794.26) | **360.60** (219.82 – 591.54) | **508.63** (291.42 – 887.73) | **8.16** (1.63 -40.98) | **5333.01** (2408.10 – 11810.55) | **7106.92** (6026.83 – 8380.58) | **8.33** (1.47 – 47.19) | **6270.33** (5308.34 – 7406.65) | **4788.66** (3101.50 – 7393.59) |
| # of SPR (NAb ≥ 1: 32) | 3 | 24 | 21 | 23 | 1 | 19 | 23 | 1 | 22 | 24 |
| Seroprotection rate (95% CI**) | **13.04** (2.78 – 33.59) | **100.00** (85.76 – 100.00) | **100.00** (83.89 – 100.00) | **100.00** (85.18 – 100.00) | **10.00** (0.25 – 44.50) | **95.00** (75.13 – 99.87) | **100.00** (85.18 – 100.00) | **12.50** (0.32 – 52.65) | **100.00** (84.56 – 100.00) | **100.00** (85.75 – 100.00) |
|  | **2 years after 2^nd^ dose (Day 759)** | | | | **1 year after booster dose (Day 731)** | | | **1 year after booster dose (Day 731)** | | |
| N | 23 | 24 | 21 | 23 | 10 | 20 | 23 | 8 | 22 | 24 |
| GMT (95% CI*) | **6.51** (3.66 – 11.56) | **180.02** (110.23 – 293.99) | **267.49** (156.61 – 456.89) | **375.68** (232.85 – 606.13) | **4.0** (4.0 – 4.0) | **1490.24** (1023.84 – 2169.08) | **1421.22** (943.92 – 2139.86) | **7.64** (1.65 – 35.29) | **930.67** (540.98 – 1601.09) | **1196.47** (668.74 – 2140.64) |
| # of SPR (NAb ≥ 1: 32) | 3 | 23 | 21 | 23 | 0 | 20 | 23 | 1 | 22 | 24 |
| Seroprotection rate (95% CI**) | **13.04** (2.78 – 33.59) | **95.83** (78.88 – 99.89) | **100.00** (83.89 – 100.00) | **100.00** (85.18 – 100.00) | **0.00** (0.00 – 30.85) | **100.00** (83.16 – 100.00) | **100.00** (85.18 – 100.00) | **12.50** (0.32 – 52.65) | **100.00** (84.56 – 100.00) | **100.00** (85.75 – 100.00) |

*Two-sample t test

**Binomial distribution estimation

Table S2. Safety physical examination (ITF population)

|  | **Group 2b** | | | | **Group 2c** | | | **Group 2d** | | |
| --- | --- | --- | --- | --- | --- | --- | --- | --- | --- | --- |
|  | **Placebo** | **LD** | **MD** | **HD** | **Placebo** | **MD** | **HD** | **Placebo** | **MD** | **HD** |
| **Year 3 (Day 1096)** |  | | | |  | | |  | | |
| **N** | - | - | - | - | **12** | **31** | **33** | **8** | **26** | **26** |
| **At least one of below** | - | - | - | - | **2** | **1** | **5** | **2** | **0** | **0** |
| Chest |  |  |  |  | 1 | 0 | 0 | 0 | 0 | 0 |
| Eyes, ENT, Mouth & Tongue |  |  |  |  | 2 | 0 | 3 | 1 | 0 | 0 |
| Respiratory |  |  |  |  | 2 | 1 | 1 | 1 | 0 | 0 |
| Skin, Nails & Hair |  |  |  |  | 0 | 0 | 1 | 1 | 0 | 0 |
| **Year 4 (Day 1461)** |  | | | |  | | |  | | |
| **N** | **22** | **23** | **24** | **22** | **12** | **31** | **32** | **7** | **24** | **25** |
| **At least one of below** | **1** | **0** | **0** | **0** | **0** | **2** | **1** | **0** | **0** | **0** |
| Chest | 0 | 0 | 0 | 0 | 0 | 1 | 0 | 0 | 0 | 0 |
| Eyes, ENT, Mouth & Tongue | 1 | 0 | 0 | 0 | 0 | 1 | 0 | 0 | 0 | 0 |
| Respiratory | 0 | 0 | 0 | 0 | 0 | 0 | 1 | 0 | 0 | 0 |
| Skin, Nails & Hair | 0 | 0 | 0 | 0 | 0 | 0 | 0 | 0 | 0 | 0 |
| **Year 5 (Day 1826)** |  | | | |  | | |  | | |
| **N** | **21** | **23** | **23** | **22** | **12** | **30** | **33** | **7** | **24** | **24** |
| **At least one of below** | **1** | **0** | **1** | **1** | **0** | **0** | **0** | **0** | **0** | **0** |
| Chest | 1 | 0 | 0 | 0 | 0 | 0 | 0 | 0 | 0 | 0 |
| Eyes, ENT, Mouth & Tongue | 0 | 0 | 1 | 0 | 0 | 0 | 0 | 0 | 0 | 0 |
| Respiratory | 0 | 0 | 0 | 0 | 0 | 0 | 0 | 0 | 0 | 0 |
| Skin, Nails & Hair | 0 | 0 | 0 | 1 | 0 | 0 | 0 | 0 | 0 | 0 |
